# A structural mean modelling Mendelian randomization approach to investigate the lifecourse effect of adiposity: applied and methodological considerations

**DOI:** 10.1101/2024.03.27.24304961

**Authors:** Grace M. Power, Tom Palmer, Nicole Warrington, Jon Heron, Tom G. Richardson, Vanesa Didelez, Kate Tilling, George Davey Smith, Eleanor Sanderson

## Abstract

The application of a lifecourse approach to genetic epidemiology is key to better understanding causal effects of adversities on health outcomes over time. For some time-varying phenotypes, it has been shown that genetic effects may have differential importance in the development of an exposure at different periods in the lifecourse. Mendelian randomization (MR) is a technique that uses genetic variation to address causal questions about how modifiable exposures influence health. MR studies often employ conventional instrumental variable (IV) methods designed to estimate lifelong effects. Recently, several extensions of MR have been used to investigate time-varying effects, including structural mean models (SMMs). SMMs exploit IVs through g-estimation and circumvent some of the parametric assumptions of other MR methods.

In this study, we apply g-estimation of SMMs to MR. We aim to estimate the period effects of adiposity measured at two different life stages on cardiovascular disease (CVD), type 2 diabetes (T2D) and breast cancer in later life. We found persistent period effects of higher adulthood adiposity on increased risk of CVD and T2D. Higher childhood adiposity had a protective period effect on breast cancer. We compare this method to an inverse variance weighted multivariable MR approach: a technique also using multiple IVs to assess time-varying effects, however, relying on a different set of assumptions and subsequent interpretations. We discuss the strengths and limitations of each approach and emphasise the importance of underlying methodological assumptions in the application of MR to lifecourse research questions.

## Introduction

Exposures throughout life (including *in utero)* set in place the structures that shape future health outcomes (1). A lifecourse approach investigates the contribution of early and later life exposures to identify risk and protective mechanisms across the lifespan (2-4). Confounding, in particular time-modified confounding (when the causal relation between a confounder and the exposure or outcome changes over time), time-varying confounding (where earlier levels of the exposure causally affect later values of the confounder of future measures of the exposure) and intermediate confounding (a confounder of the mediator-outcome relationship), is anticipated in studies with earlier life and time-varying exposures and later life health outcomes. Mendelian randomization (MR) exploits the random assortment of genetic variants, independent of other traits, to enable analyses that largely mitigate against distortions resulting from confounding bias and reverse causality (6). Within a causal inference framework, MR builds on instrumental variable (IV) analysis, whereby specific conditions are required to hold (7). These are that the IV must (i) be associated with the exposure of interest (‘relevance’), (ii) not share common causes with the outcome (‘independence’) and (iii) not affect the outcome other than through the exposure (‘exclusion-restriction’) (8, 9).

For some exposures, it has been shown that genetic effects may have differential importance in the development of an exposure at different periods in the lifecourse (10-14). MR studies, however, typically use a single exposure measurement to estimate the ‘total’ effect of an exposure on an outcome. As such, effect estimates for a single measure of a time-varying exposure, such as adiposity, using “standard” MR designs, may be interpreted incorrectly if the time-varying nature of the relationship between the genetic variants and the exposure is not taken into consideration (15). MR methods that rely on having multiple instruments with different effects on an exposure at differing times have been developed, or adapted, to address lifecourse research questions (11, 16-28). These include inverse variance weighted multivariable MR (IVW-MVMR) and MR using g-estimation of structural mean models (SMM-MR) (11, 27).

IVW-MVMR estimates the controlled effects of several potentially correlated exposures on an outcome, where each effect is controlled for other exposures included in the model (11, 19). The method relies on three assumptions, which expand on those highlighted above: (i) the ‘relevance’ assumption is modified to indicate that the liability (an underlying normally distributed latent (unmeasured) variable) to each exposure is robustly predicted by the genetic variants controlled for other exposures included in the estimation, (ii) the ‘independence’ assumption remains unchanged, and (iii) the ‘exclusion-restriction’ assumption proposes genetic variants are not associated with the outcome other than via liabilities to exposures included in the estimation (11, 29).

In a lifecourse setting, the correlated exposures in IVW-MVMR correspond to repeated measures of the same exposure over time. For example, IVW-MVMR has been used to evaluate whether childhood adiposity has an effect on disease outcomes after controlling for adiposity in adulthood (19, 30-33). If the main exposure (i.e., genetically predicted childhood adiposity) has a causal influence on disease risk, IVW-MVMR allows us to decipher whether this effect is not acting through adulthood adiposity or partly mediated by adult adiposity. An earlier application of this method showed that while the total effects indicated increased childhood adiposity was a risk factor for coronary heart disease (CHD) and type 2 diabetes (T2D), the controlled ‘direct’ effect estimates for childhood adiposity after controlling for later life adiposity were compatible with, and close to, the null (19). This suggests that the influences of childhood adiposity on these outcomes are mediated by adiposity in adulthood, highlighting an important public health message: that adverse effects may be minimised or reversed by preventing obesity before adulthood (34). Findings from this study additionally indicated that increased adiposity in childhood had a protective controlled effect on breast cancer risk with less evidence of an adult effect, suggesting that the effect of childhood adiposity is unlikely to be mediated by adulthood adiposity. Prostate cancer was also investigated, though there was very little evidence of a causal effect of either early or later life measures on this outcome (19).

An alternative MR approach to investigate time-specific effects has been proposed through the application of SMMs (27), which exploit IVs through g-estimation (35, 36). SMMs are semi-parametric, circumventing some parametric assumptions made by other MR methods such as the assumption of a linear relationship with a continuous outcome when using two-stage least-squares (2SLS) (37). SMMs comprise a wide array of models, including those with binary, categorical and time-to-event outcomes.

Three causal parameters, described previously by Shi *et al.* (27), can be identified using a SMM approach if combined with suitable IVs satisfying specific assumptions: a point effect, a period effect, and a lifetime effect. A point effect is the effect of raising an exposure by a unit at only one precise single time point on the outcome. A period effect is the effect of a unit increase in the exposure on the outcome over a specified period of time. This period effect can be defined as the difference between the mean counterfactual outcome had everyone received a particular exposure and the mean counterfactual outcome had everyone received an exposure a unit higher between times *m* − *p* and *m* (where *m* = age at end of time period and *p* = length of time period). Finally, a lifetime effect is the effect of a unit increase across the entire lifecourse (defined as starting at conception and ending at the outcome recording time) (27). This is identical to a period effect where the beginning and end of the period have been defined to cover the lifetime.

Depending on the plausibility of assumptions, it is not a requirement to have multiple measures for an exposure to estimate any of the effects defined. To estimate a point effect the genetic variants used as an IV needs to have a direct effect on the exposure only at the time point considered and not at any other time point. This is unlikely to be plausible in many settings. Period or lifetime effects can be estimated with one measure of the exposure if the IV has the same effect on the exposure throughout the whole period, and no direct effect on the exposure outside that period. In our data application, we consider the effect of adiposity measured around age 10 on a set of adulthood outcomes. To interpret this as a point effect we would assume that the instrument used directly effects adiposity at age 10 only. To interpret this as a period effect, we might assume that our measure of adiposity represents the preadolescent period (∼9-12 years) as opposed to age 10 specifically and that our instrument has a constant effect on the exposure over this period and no direct effect at any other age. To interpret a lifetime effect when using a single exposure measurement, we would assume that the instrument has the same effect on the exposure across the lifecourse up to the point the outcome is measured.

Shi *et al.* show that to satisfy the final instrumental condition aligned with those pertaining to IVW-MVMR described earlier (i.e., that genetic variants are not associated with the outcome other than via liabilities to exposures included in the estimation), each component of the time-varying exposure outside of the period considered must not be directly affected by the instrument or not directly affect the outcome. Under this assumption, using an example with an exposure measured at one time period (*X****_2_***), when investigating this effect on *Y*, there should be no direct pathway from genetic variants (*Z*) to *X****_1_*** and *X****_3_*** or no pathway from *X****_1_*** or *X****_3_*** directly to *Y* (see Figure 1). Presence of such pathways would induce horizontal pleiotropy. If multiple time periods had been measured and included in this model, the same assumptions would apply.

**Figure 1.**
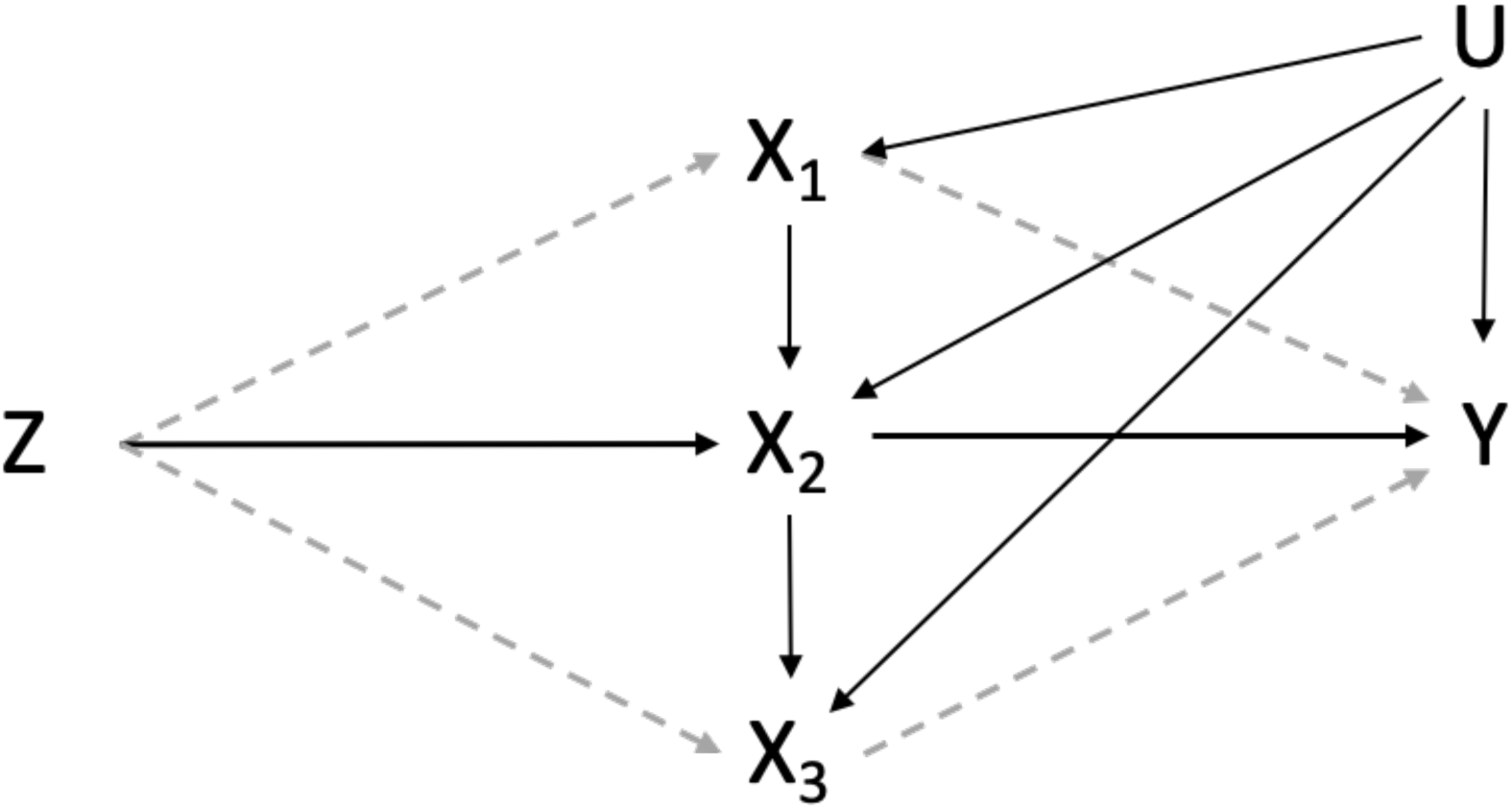
Causal diagram for instrumental variable analyses with one measured period (X2) (using instruments associated with three potential time periods) of an exposure. *Z* is a set of genetic variants associated with X2, *Y* is an outcome observed at one time only, X**_1_** and X**_3_** indicate potentially relevant periods that are unmeasured. U is a set of unobserved confounders of the exposure at each time period and the outcome. X**_1_**, X2, X**_3_** and *Y* are confounded by a set of unobserved confounders U. Dashed lines indicate paths via unmeasured exposures.

Childhood adiposity strongly influences adulthood adiposity (38). Genetic variants with varying levels of association with childhood and adult adiposity may be targeting different causal pathways underlying the relationship between adiposity and complex outcomes, that vary in importance at different life stages (39). For example, there have been situations where genetic variants associated with adulthood adiposity may have predicted a component of adiposity in early life not detected by childhood associated genetic variants (39). This can lead to deviations from the gene-environment equivalence assumption in MR (40), which asserts that the effect of germline genetic perturbations should have the same downstream consequence on outcomes as if they were caused by the modifiable exposures themselves (39). This is important to consider when selecting genetic variants to instrument period effects as described.

An alternative interpretation that has been proposed using IVW-MVMR is that there may be a path from the instruments to the outcome via exposures at other periods which have not been included in the estimation and this path will form some of the effect estimate obtained (11, 21). Consequently, the estimand being targeted is the effect of an increase in one unit of liability to the exposure. With reference to Figure 2, the instruments associated with the liability (*L*) in at least one period of the lifecourse may be associated with *L* in different periods. In addition, the degree of this association may vary across periods. The genetically predicted effect of increasing the exposure by a unit of liability at a particular time period will thus include genetic effects on the exposure at other time periods if the exposure in those periods has an effect on the outcome. If each genetic variant has a different trajectory for the liability, then the effects estimated will be an average across these.

**Figure 2.**
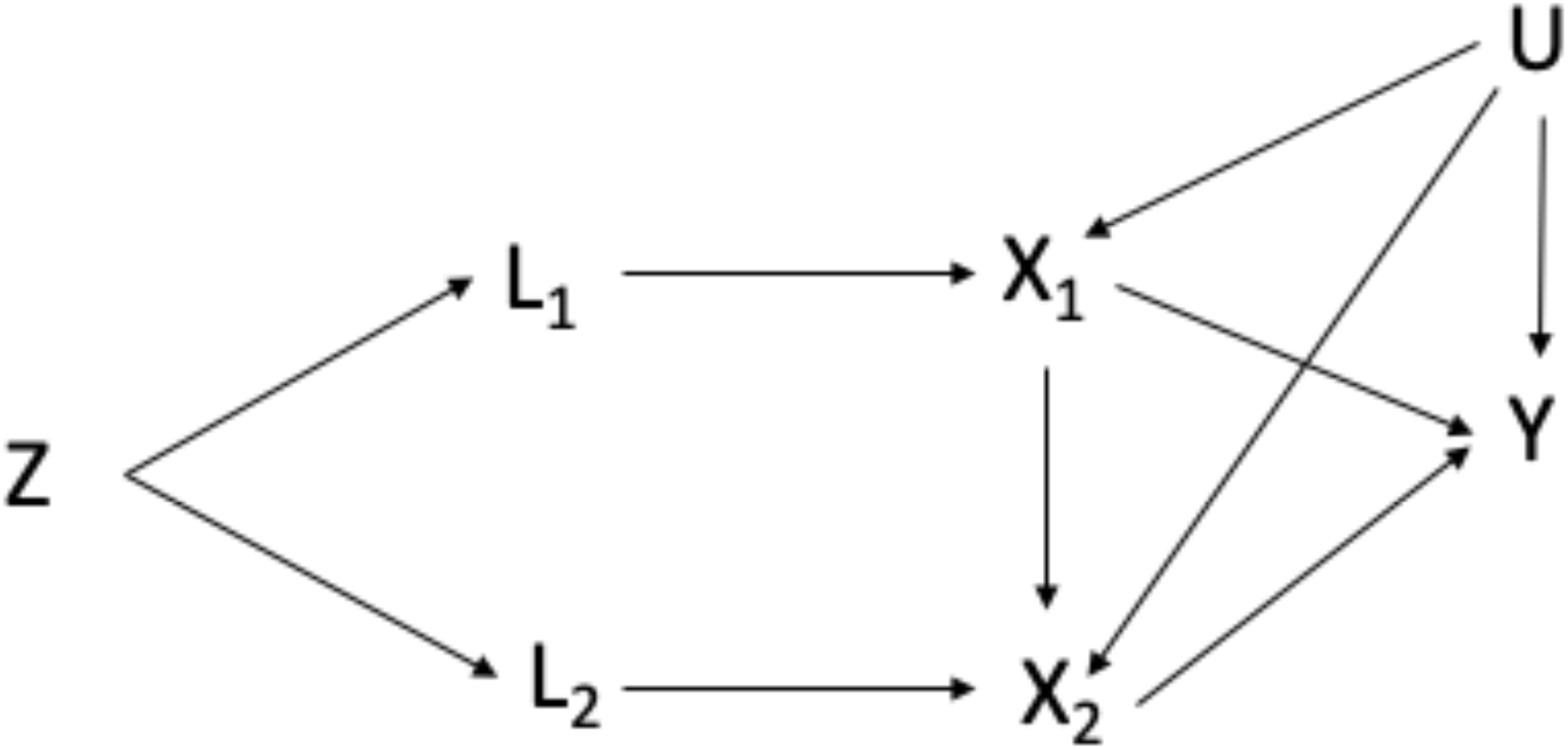
Causal diagram for instrumental variable analyses with two periods (using instruments associated with two time points assumed to represent two respective periods) of exposure (adapted from Sanderson et al. (11)). L1 is liability to the exposure in the earlier period, L2 is liability to the exposure in the later time period, is a set of genetic variants associated with L1 and L2. X1 is a measure of the exposure in the early time period, X2 is a measure of the exposure at the second time period, *Y* is an outcome observed at one time only, U is a set of unobserved confounders of the exposure at each time period and the outcome. We have assumed that there is no time-varying confounding.

Throughout we assume no time varying confounding, which is where earlier levels of the exposure causally affect later values of the confounder. Such confounding creates a feedback loop over time between the exposure and the confounder as later values of the exposure will then be affected by the confounder. When conducting MR to answer lifecourse research questions, these feedback loops are part of the causal pathway that we are estimating as they are part of the effect of that exposure on the outcome (Figure 3).

**Figure 3.**
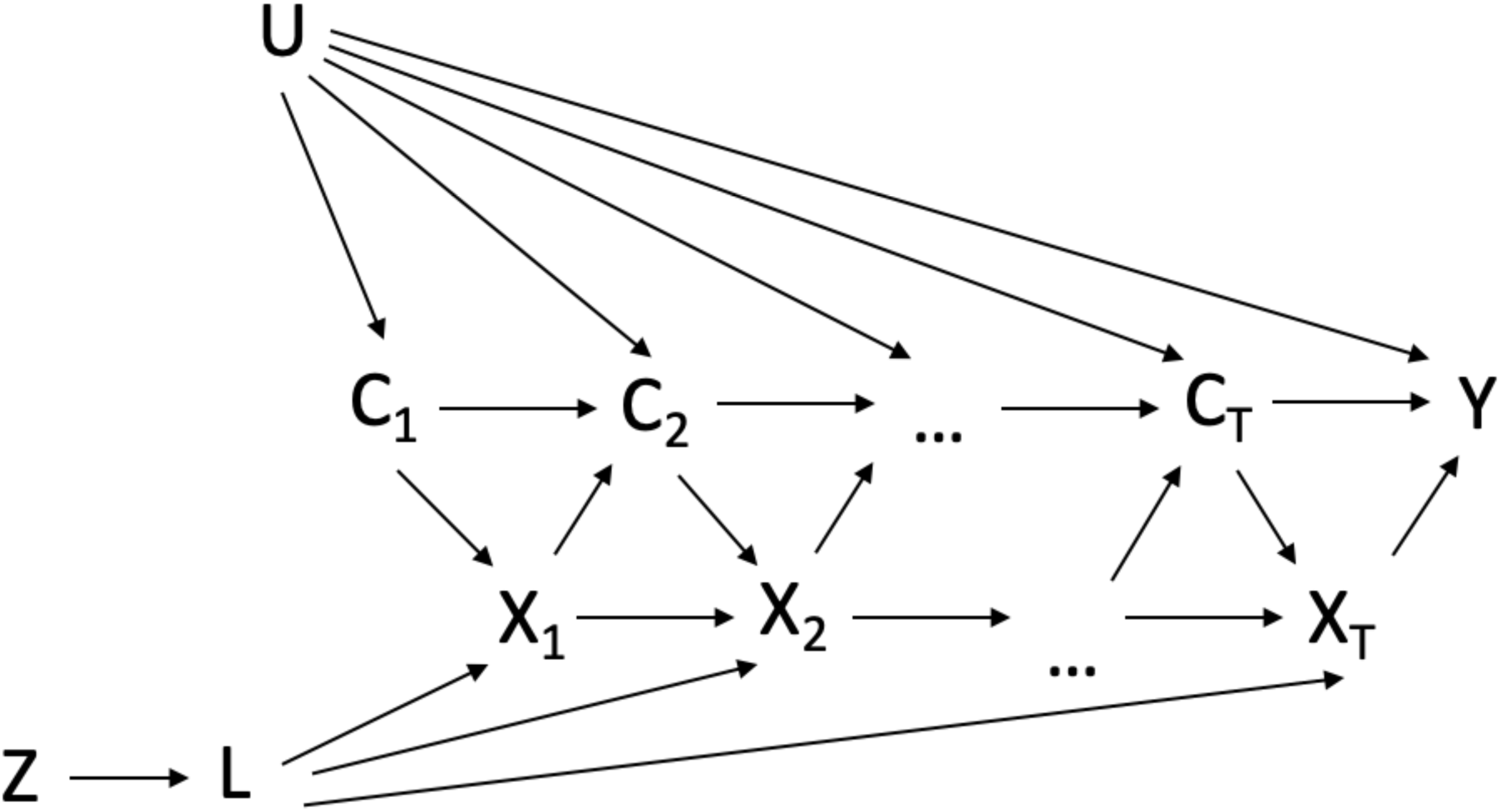
Causal diagram depicting time-varying confounders {C_1_, C_2_,…,C_T_} affected by exposure (X), when the outcome (Y) is measured at the end of follow-up. {X_1_, X_2_,…,X_T_} represent the exposure variables of interest measured at time points 1, 2,…,T. {C_1_, C_2_,…,C_T_} represent the potential confounders measured at time points 1, 2,…,T, where it is assumed that C_T_ occurs just before X_T_. In this diagram, Y is the outcome of interest, measured at the end of the study (at visit T + 1), and U is a set of unmeasured factors that influence {C_1_, C_2_,…, C_T_} and Y (adapted from Daniel et al. (41)). L is the underlying liability to the exposure {X_1_, X_2_,…,X_T_}. *Z* is a set of genetic variants associated with L. The arrows in this diagram represent the assumed direction of causal influence. For clarity, we have not included all arrows between U and {X_1_, X_2_,…,X_T_} and {X_1_, X_2_,…,X_T_} and Y.

In this paper we apply MR using g-estimation of SMMs to estimate the period effects of measures of childhood and adult adiposity on CVD, T2D and breast cancer in later life. We use this application to illustrate and describe this approach and to provide a comprehensive interpretation of results that can be obtained. We compare results using SMM-MR to results using the same data in an IVW-MVMR framework. Finally, we compare these findings to those produced using the same instruments but different outcome data to estimate potential biases that may have resulted from sample overlap or selection (11, 19). We then discuss plausible next steps for MR methods used to address lifecourse epidemiology questions.

## Methods

### Data sources

UK Biobank data were collected between 2006 and 2010 on individuals aged between 40 and 69 years old at baseline, from clinical examinations, assays of biological samples, detailed information on self-reported health characteristics, and genome-wide genotyping, using a prospective cohort study design (48). The childhood adiposity measure applied in this study, used recall questionnaire data, involving responses from adult participants who were asked whether, compared to the average, they were ‘thinner’, ‘about average’ or ‘plumper’, when they were aged 10 years old. The adult adiposity variable was derived using clinically measured body mass index (BMI) data (mean age 56.5 years). It was then separated into a 3-tier variable based on the same proportions as the early life adiposity variable (i.e., thinner, plumper, and about average) (49, 50). This was to ensure that derived effect estimates from subsequent analyses were as comparable as possible. Individuals that did not have data for both childhood and adult adiposity were excluded from analyses. The UK Biobank study have obtained ethics approval from the Research Ethics Committee (REC; approval number: 11/NW/0382) and informed consent from all participants enrolled in UK Biobank. Estimates were derived using data from the UK Biobank (app #76538).

Genetic variants strongly associated with childhood and adult adiposity (using P < 5×10^-8^ and r^2^ < 0.001) were identified in a large-scale genome-wide association study (GWAS), previously undertaken in the UK Biobank study on 463 005 individuals, adjusting for age, sex, and genotyping chip (48, 51). The genetic instruments derived from these measures have been independently validated in two distinct cohorts, providing verification they can reliably measure childhood and adult adiposity (19, 52). These genetic variants were used to generate polygenic risk scores (PRSs) for individual-level MR analyses.

Phenotypic data for outcomes under investigation: CVD, T2D, and breast cancer, were obtained from the UK Biobank. These outcomes were classified using International Classification of Diseases, Tenth Revision (ICD-10) codes. CVD was defined as cardiovascular disease cases (ICD-10 code I210-I229, I231-36, I238, I240, I241, I248-I256, I258, I259). T2D was defined as T2D mellitus (ICD-10 code E110-19). Breast cancer was defined as malignant neoplasm of the breast (ICD-10 code C500-06, C508-09).

Genetic IVs in a MR setting are conventionally selected from an independent dataset where the sample does not overlap with the dataset being analysed (53). This is to avoid overfitting bias. Conducting MR using overlapping sets of participant samples may bias in the direction of the conventional observational results between the exposure and outcome (54). In our investigation it was not possible to avoid this. However, we compare our MVMR-IVW results to the same analysis using non-overlapping outcome data, replicating analyses previously conducted (19), to assess how using these data may have affected our results. For this analysis GWAS data for CHD, T2D and breast cancer were obtained from large scale consortia, which did not include data from the UK Biobank (55-57). Details on these datasets have been described previously (19).

### Statistical analysis

First, we employed a SMM technique in a univariable framework using polygenic risk scores (PRSs) generated from selected genetic variants to estimate period effects of childhood and adulthood adiposity on CVD, T2D and breast cancer. We then applied SMMs in a multivariable framework to compare with an IVW-MVMR approach used to estimate controlled period effects. For comparative purposes, we employed the IVW method to conduct MR analyses using summary statistics generated with the same full sets of genetic variants used in the SMM analyses. These steps are described in further detail below:

### Polygenic Risk Scores (PRS)

We used PLINK 2.00 to calculate PRSs for each individual in the study (58). We generated the PRSs using different single nucleotide polymorphisms (SNPs) association stringency thresholds with the exposure phenotypes, childhood and adulthood adiposity. The criteria for each of the PRSs are outlined in Table 1. Analyses were undertaken three times: in all eligible participants and then for each sex separately. The lists of SNPs used for each PRS can be found in Supplementary 1.

**Table 1.** Details of the SNP association thresholds used for a Mendelian randomization (MR) analysis.

| Exposure | SNP threshold criteria | Stringency rating |
| --- | --- | --- |
| Childhood adiposity | The genetic variants strongly associated* with childhood adiposity, regardless of their association with adulthood adiposity | Low |
| Adulthood adiposity | The genetic variants strongly associated* with adulthood adiposity, regardless of their association with childhood adiposity | Low |
| Childhood adiposity | The genetic variants strongly associated* with childhood adiposity and exclude SNPs that are associated with adulthood adiposity at $P \leq 5 \times 10^{-8}$ | Low-Medium |
| Adulthood adiposity | The genetic variants strongly associated* with adulthood adiposity and exclude SNPs that are associated with childhood adiposity at $P \leq 5 \times 10^{-8}$ | Low-Medium |
| Childhood adiposity | The genetic variants strongly associated* with childhood and exclude SNPs associated with adulthood adiposity at $P \leq 0.05$ with Bonferroni correction** | Medium-High |
| Adulthood adiposity | The genetic variants strongly associated* with adulthood adiposity and exclude SNPs associated with childhood adiposity at $P \leq 0.05$ with Bonferroni correction** | Medium-High |
| Childhood adiposity | The genetic variants strongly associated* with childhood adiposity and exclude SNPs associated with adulthood adiposity at $P \leq 0.05$ | High |

| Adulthood adiposity | The genetic variants strongly associated* with adulthood adiposity and exclude SNPs associated with childhood adiposity at $P \leq 0.05$ | High |
| --- | --- | --- |

\* Strongly associated refers to associated at genome wide significance ( $P \leq 5 \times 10^{-8}$ )
\*\* Genetic variants strongly associated with childhood/adulthood adiposity at genome wide significance and not adulthood/childhood adiposity (exclude child SNPs at $P \leq 0.05$ ) were counted. We then divided 0.05 by this number to generate the Bonferroni corrected P value.

### Mendelian randomization using structural mean modelling

In this study, we used individual-level data to perform MR using SMMs. We interpret results obtained as period effects. The estimation of a point effect is implausible in this context as it assumes any genetic variants would only be associated with adiposity at a single point in the lifecourse only. In addition, a lifetime effect would assume the genetic variant-exposure relationship was the same across the lifetime, however, previous work has shown the specificity of genetic variants associated with adiposity to specific periods in the lifecourse (19).

To highlight results obtained in most “standard” MR analyses we first employed a SMM technique in a univariable framework using the genetic variants strongly associated with childhood adiposity around age 10 (P≤ 5×10^-8^), regardless of their association with adulthood adiposity and the full set of genetic variants strongly associated with adulthood adiposity around 57, regardless of their association with childhood adiposity.

Next, we used the genetic variants strongly associated with childhood adiposity (P≤ 5×10^-8^), excluding SNPs strongly associated with adulthood adiposity at P ≤ 5×10^-8^, to estimate the effects of childhood adiposity for a period that started in childhood and ended at some point between 10 and 57 years. We used the genetic variants strongly associated with adulthood adiposity (P≤ 5×10^-8^), excluding SNPs associated with childhood adiposity at P ≤ 5×10^-8^, to estimate the effects of adulthood adiposity for a period that started at some point between 10 and 57 years up to the point the outcome is measured. This is attempting to estimate a childhood period effect without the effect of adulthood adiposity and an adult period effect without the effect of childhood adiposity. Definition of these periods is imprecise as the instruments and our understanding of their relationship to the phenotypic information available is limited.

To further explore the impact of instrument selection when estimating period effects as described above, we applied the other SNP association thresholds outlined in Table 1 and ran models with the PRS generated from each of these, in turn.

We additionally applied SMMs in a multivariable framework to calculate the effects of childhood adiposity and adulthood adiposity, simultaneously, on CVD, T2D and breast cancer, controlling for either adulthood adiposity or childhood adiposity, respectively. This approach estimates the controlled period effect of an exposure at a specific period by controlling for the exposure at a period not considered the main exposure period at a given value. The MR analyses conducted and their interpretation, along with the closest equivalent IVW estimator, are summarised in Table 2.

**Table 2.** Presentation of the constituent components of structural mean modelling (SMM) Mendelian randomization (MR), with the comparative interpretations and assumptions made when using inverse variance weighted (IVW) MR.

| SMM/IVW Effect | Alternative MR estimator | Exposure | Variables controlled for | Methodological assumptions being made using SMM-MR | Interpretation of the estimand (under assumptions) identified using SMM-MR | Methodological assumptions being made using IVW-MR | Interpretation of the estimand (under assumptions) identified using IVW-MR |
| --- | --- | --- | --- | --- | --- | --- | --- |
| Point | Univariable MR | Body size at a specific time point | -- | Genetic variants strongly associated with body size at a specific time point that do not have an effect through any other time point. | The effect of body size at one specific time point on the outcome | --* | --* |
| Period (varies depending on different SNP association stringency thresholds) / Total period | Univariable MR | Childhood body size | -- | Genetic variants strongly associated with childhood body size do not have an effect through another time period, such as adulthood body size. | The effect of childhood body size around the period childhood body size was measured, independent of other time periods | Genetic variants strongly associated with childhood body size may have an effect through another time period, such as adulthood body size. Genetic variants have different effects at different time periods | The effect of liability to childhood body size at around the age childhood body size was measured |
| Period (varies depending on different SNP association stringency thresholds) / Total period | Univariable MR | Adulthood body size | -- | Genetic variants strongly associated with adulthood body size do not have an effect through another time period, such as childhood body size. | The effect of adulthood body size around the period adulthood body size was measured, independent of other time periods | Genetic variants strongly associated with adulthood body size may have an effect through another time period, such as childhood body size. Genetic variants have different effects at different time periods | The effect of liability to adulthood body size at around the age adulthood body size was measured |
| Controlled period | Multivariable MR | Childhood body size | Adulthood body size | Genetic variants have different effects at the different time periods considered. | The controlled effect of childhood body size including pathways comprising body size measures from different timeframes in the lifecourse, controlled for body size measures from other time periods included | Genetic variants have different effects at different time periods | The controlled period effect of liability to childhood body size including pathways comprising body size measures from different timeframes in the lifecourse, controlled for body size measures from other time periods included |
| Controlled period | Multivariable MR | Adulthood body size | Childhood body size | Genetic variants have different effects at the different time periods considered | The controlled effect of adulthood body size including pathways comprising body size measures from different timeframes in the lifecourse, controlled for considered body size measures from other time periods | Genetic variants have different effects at different time periods | The controlled period effect of liability to adulthood body size including pathways comprising body size measures from different timeframes in the lifecourse, controlled for considered body size measures from other time periods |
| Lifetime | Univariable MR | Body size | -- | Relationship between genetic variants and body size are constant for body size over the lifecourse | The effect of body size over the entire lifecourse on an outcome | -- | The effect of liability to body size over the entire lifecourse on an outcome |

\*Estimation of the point effect IVW-MR is technically plausible; however, I do not recommend this. Assumptions and interpretation would be the same as with SMM-MR. IVW: Inverse variance weighted; MR: Mendelian randomisation; SNP: Single-nucleotide polymorphism.

### Genome-wide Association Studies (GWAS)

We used BOLT-LMM to assess the association between genetic variants across the human genome on the three outcomes of interest: CVD, T2D and breast cancer. The results from these GWAS were used to run the IVW-MR analyses (59, 60). BOLT-LMM applies a Bayesian linear mixed model to estimate the association between each genetic variant and each of the outcome measures, while accounting for both relatedness and population stratification. We added age at baseline, sex, and type of genotyping array as covariates in the model.

### Mendelian randomization using inverse variance weighting

We employed the inverse variance weighted method (IVW) to conduct further MR analyses IVW takes SNP-outcome estimates and regresses them on the SNP-exposure associations We ran IVW-MR on outcomes from two sets of data sources. First, we used the GWAS generated from UK Biobank data to compare against the MR analyses using SMMs. Second, we used the GWAS obtained from large scale consortia, which did not include data from the UK Biobank (55-57) used in a previous study (19), to assess how using overlapping outcome data may have affected our results.

For each of the sets of IVW analyses, we estimated the ‘total’ period effect estimates between genetically predicted childhood and adulthood adiposity separately with each of the disease outcomes of interest in a univariable framework. We then ran MVMR analyses to estimate the controlled period effect of childhood adiposity on each outcome in turn, controlling for adult adiposity, as well as the effect of adult adiposity on each outcome, controlling for childhood adiposity.

The IVW-MR analyses were conducted using the TwoSampleMR R package (63). Plots in this paper were generated using the R package ‘ggplot2’ (64). These analyses were undertaken using R (version 3.5.1).

## Results

Univariable SMM-MR analyses indicated evidence of an effect of higher childhood adiposity (regardless of SNP association with adulthood adiposity) on increased CVD and T2D risk (estimated change in the effects of the probability of the outcome for a unit change in adiposity i.e., risk difference (RD)), 95% CI: 0.021, 0.013 to 0.029 and 0.049, 0.043 to 0.054, respectively) as well as of higher adult adiposity on increased risk of CVD and T2D (RD, 95% CI: 0.062, 0.056 to 0.068 and 0.107, 0.101 to 0.113, respectively) (Figure 4 Supplementary 2A and 2B).There was very little evidence indicating period effects of higher childhood adiposity (excluding SNPs associated with adulthood adiposity at P≤5×10^-8^) on increased CVD risk and some evidence on the risk of T2D (RD, 95% CI: 0.004, -0.008 to 0.016 and 0.018, 0.008 to 0.028, respectively). There was strong evidence of a period effect of higher adult adiposity (excluding SNPs associated with childhood adiposity at P ≤5×10^-8^) on increased CVD and T2D risk (RD, 95% CI: 0.067, 0.059 to 0.075 and 0.109, 0.103 to 0.115, respectively).

**Figure 4.**
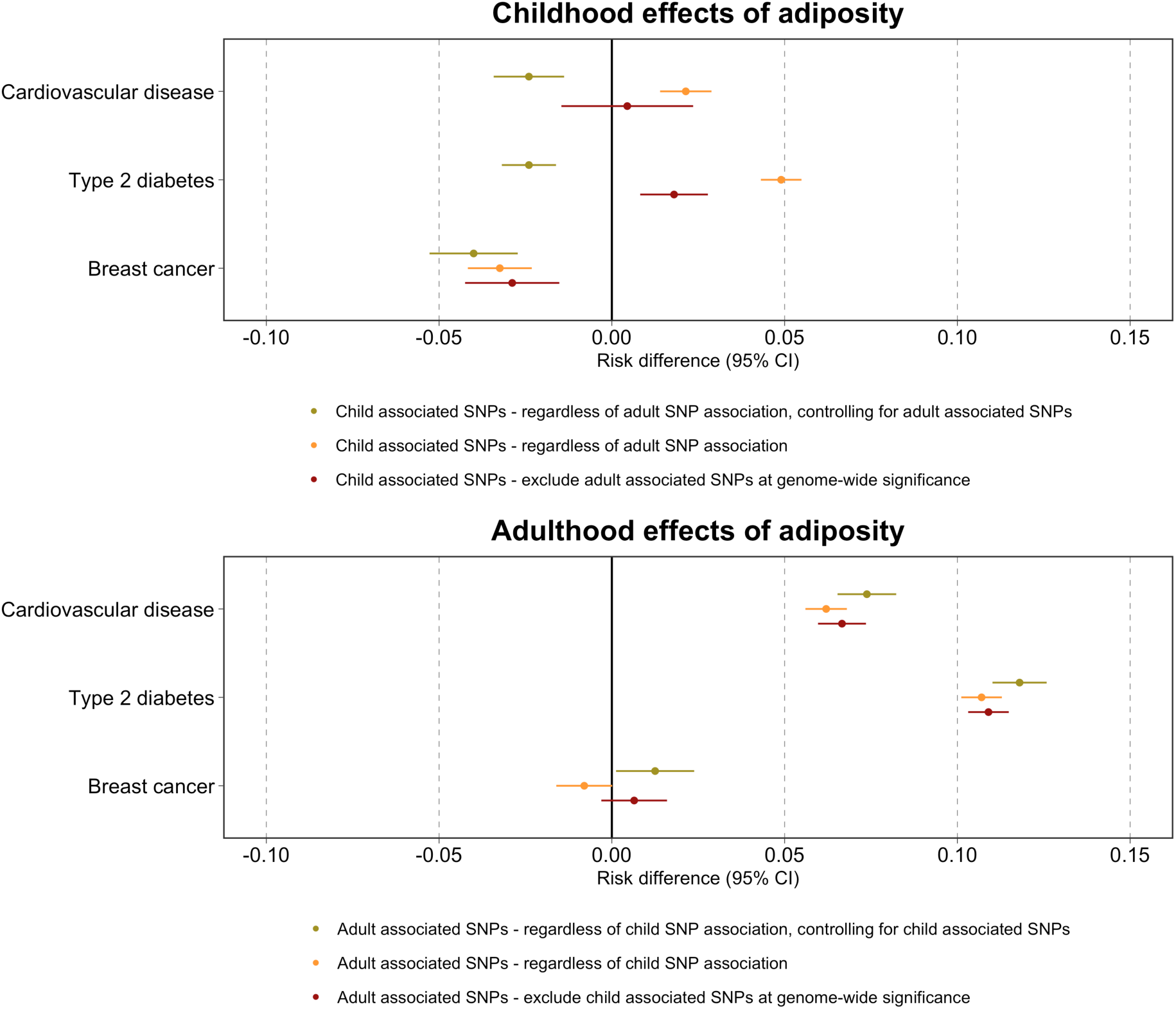
Causal risk difference estimates from univariable and multivariable MR using structural mean models (SMMs) for childhood and adulthood adiposity period effects on the outcome measures listed. Associated SNPs refers to strongly associated at genome wide significance (P≤ 5×10^-8^).

SMM-MVMR analyses showed a period effect of higher childhood adiposity on reduced CVD and T2D risk, after controlling for adult adiposity (RD, 95% CI: -0.024, -0.034 to -0.014, and -0.024, -0.032 to -0.016, respectively). There was a period effect of higher adult adiposity on the CVD and T2D risk in later life, after controlling for childhood adiposity (RD, 95% CI: 0.074, 0.066 to 0.082 and 0.118, 0.110 to 0.126, respectively).

Univariable SMM-MR analyses indicated evidence of an effect of higher childhood adiposity (regardless of SNP association with adulthood adiposity) on reduced risk of breast cancer (RD, 95% CI: -0.032, -0.042 to -0.022) (Figure 4, Supplementary 2C). There was evidence of a period effect of higher childhood adiposity (excluding SNPs associated with adulthood adiposity at P≤5×10^-8^) on reduced breast cancer risk (RD, 95% CI: -0.029, -0.043 to -0.015). There was very little evidence of a period effect of higher adult adiposity (excluding SNPs associated with childhood adiposity at P≤5×10^-8^) on breast cancer (RD, 95% CI: 0.006, -0.004 to 0.016). SMM-MVMR analyses also showed evidence that higher childhood adiposity reduced the risk of breast cancer, after controlling for adult adiposity (RD, 95% CI: -0.040, -0.054 to -0.026) and that higher adult adiposity slightly increased the risk of breast cancer in later life, controlling for childhood adiposity (RD, 95% CI: 0.013, 0.001 to 0.025).

When using different SNP association stringency thresholds with the exposures to estimate period effects, univariable SMM-MR analyses indicated consistently strong evidence that higher adulthood adiposity increased risk of CVD (Figure 5A, Supplementary Table 2A) and T2D (Figure 5B, Supplementary Table 2B). When we increase instrument stringency and reduce the number of SNPs associated with adult adiposity, we see very little evidence of an effect of childhood adiposity on CVD. We see some evidence of an effect remaining in the set of SNPs with the lowest stringency rating (exclusion threshold of adulthood adiposity SNPs at P≤5×10^-8^) on T2D.

**Figure 5.**
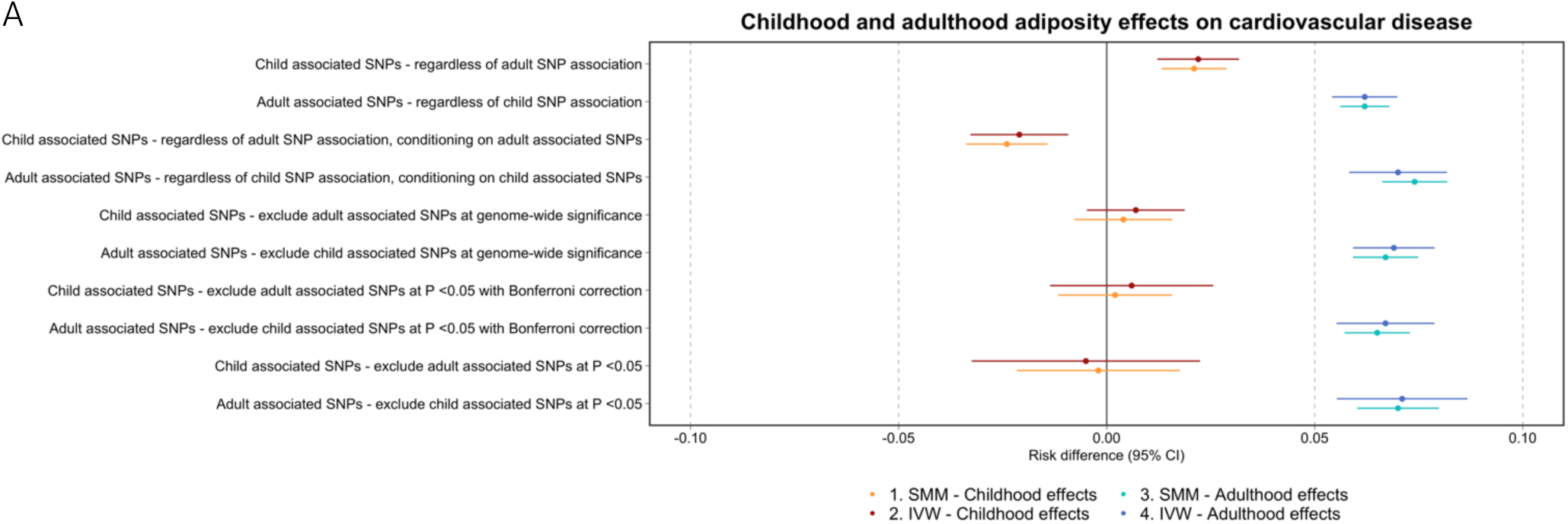
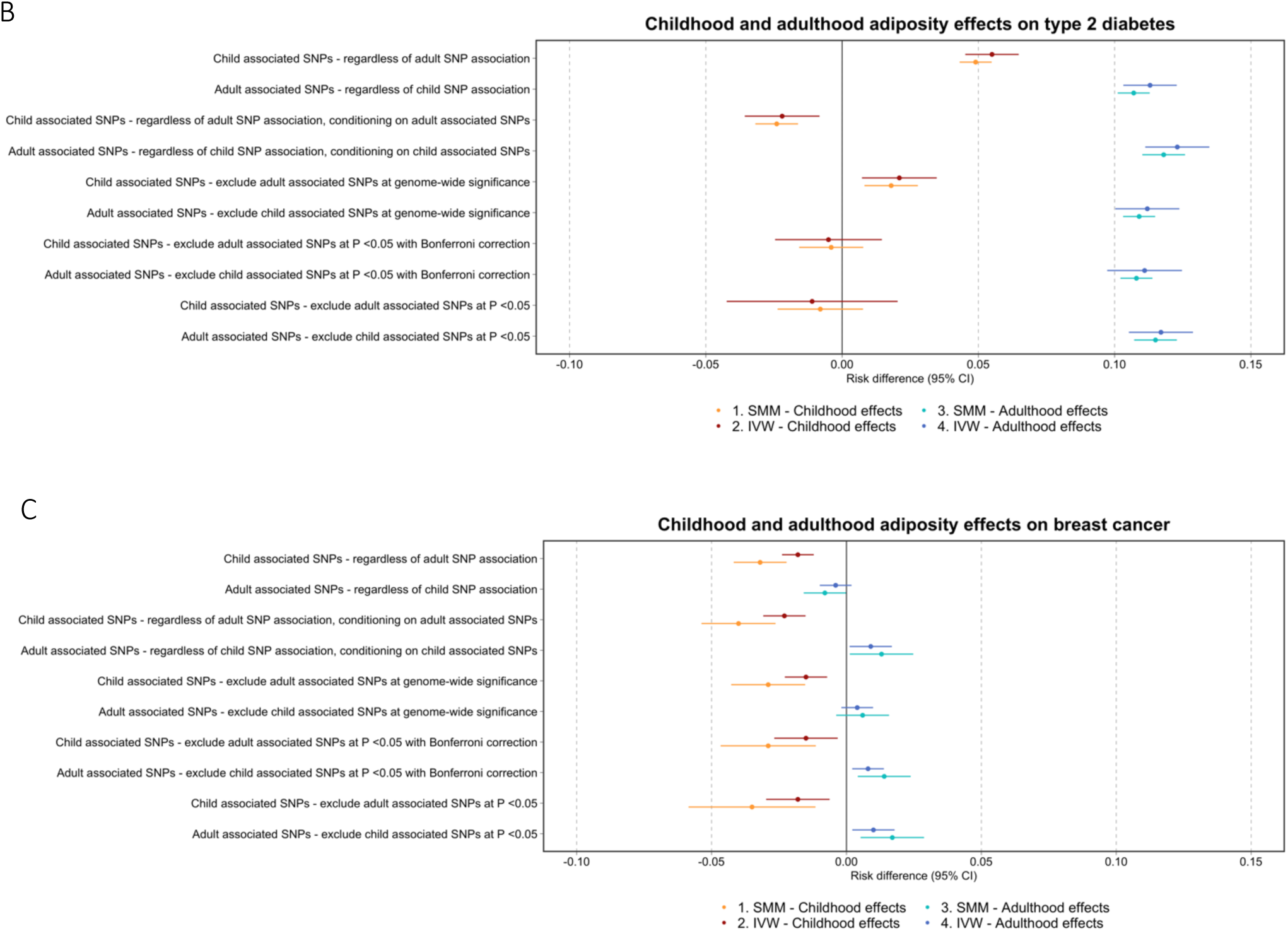
Causal risk difference estimates from univariable and multivariable MR using structural mean models (SMMs) and inverse variance weighted models (IVW) for childhood and adult adiposity and liability to childhood and adult adiposity, respectively, on the outcome measures listed. (A) Childhood and adult adiposity period effects on cardiovascular disease (CVD). (B) Childhood and adult adiposity period effects on Type 2 diabetes (T2D). (C) Childhood and adult adiposity period effects on breast cancer. Associated SNPs refers to strongly associated at genome wide significance (P≤ 5×10^-8^).

These analyses additionally indicated consistently strong evidence that higher childhood adiposity reduced the risk of breast cancer (Figure 5C, Supplementary Table 2C). A protective effect of adulthood adiposity was observed when including all SNPs regardless of association with childhood adiposity. However, when using a more stringent threshold for excluding childhood adiposity SNPs the observed protective effect of adulthood adiposity on breast cancer diminished or reversed direction.

The IVW univariable and MVMR analyses using the outcomes generated from UK Biobank data, produced results indicating very similar trends (though generally slightly weaker) to those using the SMM approach, employing the same dataset (Figure 5A-C, Supplementary Table 3A-C) The IVW univariable and MVMR analyses using the outcomes generated from the large-scale consortia indicated the same directions of effect to those using the SMM approach (Supplementary Table 4A-C, Supplementary Figure 5A-C), though with much larger effects and confidence intervals. This may be a result of Winner’s curse since there is overlap between the discovery sample and the dataset used in the core analysis (42). We found evidence that an increase in liability to adulthood adiposity increased the risk of breast cancer (Figure 5C, Supplementary Table 3C) when using UK Biobank data. We found very little evidence of an effect of adulthood adiposity on breast cancer when using the large scale meta-analysed outcome consortia data (Supplementary Table 4C, Supplementary Figure 5C).

## Discussion

In this investigation, we apply MR to lifecourse epidemiology using SMM and IVW approaches with a time-varying exposure. SMM-MR and IVW-MR target different causal estimands and key differences between these methods are their underlying assumptions and resulting interpretations of findings. In the context of this study, SMM-MR computed the effect of increasing adiposity by one unit of measurement and IVW-MR, the effect of an increase in one unit of liability to adiposity. For SMM-MR, each component of the time-varying exposure outside of the period considered must be unaffected by the instrument or affect the outcome only by affecting exposures during the time period under consideration. For IVW-MR, the genetically predicted effect of increasing the exposure by a unit of liability at one time period will include genetic effects on the exposure at other time periods if the exposure in those periods has an effect on the outcome.

Time-varying confounding occurs when earlier measures of the exposure causally affect later confounders. This confounding creates a feedback loop over time between the exposure and the confounder as later values of the exposure will become affected by the confounder. When conducting MR to answer lifecourse research questions, these feedback loops are part of the causal pathway being estimated, as they are part of the effect of that exposure on the outcome. We have assumed throughout that there is no time-varying confounding. We have also assumed all models are linear and that there are no interactions between the exposure at different time periods.

We ran analyses to estimate several period effects of adiposity on CVD, T2D and breast cancer. First, we evaluated the period effects of childhood and adult adiposity around the age of 10 and 57 years, respectively, using genetic variants strongly associated with adiposity at each period, regardless of their association with adiposity at the alternate period. We evaluated more stringently instrumented period effects of childhood (starting in childhood and ending at some point between 10 and 57 years) and adult (starting at some point between 10 and 57 years up to 57 years) adiposity. Then, we evaluated the controlled period effects of childhood and adult adiposity, controlling for either adult adiposity or childhood adiposity, respectively.

Overall, we identified persistent period effects of higher adulthood adiposity on increased risk of CVD and T2D. When instruments used more stringent SNP-association thresholds, childhood adiposity showed almost no effect on CVD and T2D. In addition, higher childhood adiposity had a protective effect on breast cancer. Higher childhood body size has been shown to be protective against breast cancer previously (363). Conversely, more stringently instrumented period as well as controlled period effects indicated that higher adulthood adiposity may increase breast cancer risk.

Our results highlight the importance of considering the potential association of the genetic variants with other time periods when estimating period effects in a SMM setting. For example, when we increased the stringency for SNP inclusion in childhood instruments, we see very little evidence of an effect of childhood adiposity on T2D. However, we see evidence of an increased total period effect and period effect in the set of SNPs strongly associated with childhood adiposity and not adulthood adiposity at P ≤ 5×10^-8^. This shows that SNPs being used to instrument adiposity at a specific period might still influence adiposity at the alternate period, just at a lower significance level resulting in them not being filtered out. Importantly, since we only have the two time periods measured in this analysis, we are not able to determine whether SNPs would associate with other time periods, outside of those measured.

As discussed previously, childhood adiposity has a causal influence on adulthood adiposity (38). By imposing a SNP selection process to conduct the SMM method we are generating and using a non-representative sample of childhood or adulthood adiposity associated SNPs. Since adiposity is a heterogeneous phenotype, SNPs associated with childhood and not adult adiposity or contrariwise may be targeting different causal pathways that vary in importance depending on the lifecourse (39) which may lead to deviations from the gene-environment equivalence assumption in MR (40). Therefore, any selection applied to SNPs for estimation of period effects should only be considered with caution.

The interpretation of a lifetime effect (which uses non-selected SNPs) aligns with the interpretation of a total effect often adopted when using IVW-MR. However, we are not able to disentangle whether earlier periods affect the outcome directly, through later periods, or if all the effect seen is due to the influence of the genetic variants used on the later periods. Equally, it is not possible to tell whether later periods affect the outcome directly, or if all the effect seen is due to the confounding influence of the earlier periods on the later period and the outcome. To allow estimation of period-specific effects under specified assumptions, we can also use a SMM- or IVW-MVMR approach, to estimate the controlled period effect of each exposure on the outcome controlled for all the other exposures included in the estimation.

The estimated results from this method will, however, still include the effects of other periods that are not included in the estimation if they are influenced by the genetic variants used to instrument the included periods. When researchers use genetic instruments that are not specific to a time period, which is commonplace in MR analyses, they are implicitly targeting a lifetime effect of liability to a unit higher level of the exposure at the time it was measured (20).

The IVW-MVMR results generated from largescale consortia indicated very little evidence of a controlled effect of adult adiposity on breast cancer (19). The risk increasing effect we observed could be due to higher adulthood adiposity influencing onset and survival from breast cancer differently in the UK Biobank cohort study compared to the case-control studies used in producing the largescale outcome data. Differences in these effects may also be partially due to a “healthy volunteer” selection bias. The UK Biobank is not representative of the sampling population - participants are a sample of volunteers with healthier lifestyles, higher levels of education and better health than the general UK population (43, 44). If participation in the UK Biobank is a consequence of either our exposure or outcome then a key MR condition (exchangeability) is violated, inducing an association between genetic instruments and unmeasured confounders of the exposure–outcome relationship (44). Another reason for inconsistencies could be weak instrument bias. This may push a null effect to be non-null, as seen in the effect between adulthood adiposity and breast cancer risk (45). In addition, when exposure and outcome effects are estimated in a single (i.e. fully overlapping) sample, as they were for our analyses, bias will be in the direction of the (confounded) observational association (46).

Shi *et al.* propose two key advantages of SMMs compared to IVW-MR (27). The first is that SMMs can be naturally extended to many settings, including accommodating failure-time outcomes and estimating effects on the multiplicative scale. The second is that SMMs are semiparametric. They therefore avoid some 2SLS parametric assumptions. In addition, Shi *et al.* claim that SMMs are more robust to model misspecification for binary outcomes (27). There is no meaningful difference between results obtained using SMM and 2SLS MR in relatively simple models such as the one we have used, however, situations in which SMMs may provide more reliable estimates for MR, specifically when addressing lifecourse questions, should be explored in future work.

Comprehensively understanding the interpretation of results after employing these methods is of key public health importance. For instance, when considering the potential differences in mediating pathways from one period (i.e., childhood adiposity) and not another (i.e., adulthood adiposity) we might hypothesise that prepubertal childhood and adult adiposity could be distinct phenotypes; an insight gained through conducting these approaches. This helps us to further unpick pathways associated with disease risk. For example, there are long run effects of childhood adiposity on physical development shown though mammographic density being a strong mediator for the protective effects of childhood adiposity on breast cancer (47).

## Conclusion

This study emphasises the importance of underlying methodological assumptions in the application of MR to lifecourse research questions and explores how output from analytical frameworks that rely on different conditions should be interpreted. It additionally highlights the careful consideration required for instrument selection when running lifecourse MR methods. We do not advocate for a particular strategy but encourage practitioners to thoroughly think through their research question, instrumental variable selection, model conditions and data availability before pursuing a particular MR approach within a lifecourse setting.

## Data Availability

This research has been conducted using the UK Biobank resource under application number 76538. All data are available within the article, the supplementary material or from the authors upon request.

## Research Ethics and Informed Consent

The UK Biobank study have obtained ethics approval from the Research Ethics Committee (REC; approval number: 11/NW/0382). The UK Biobank study have obtained informed consent from all participants enrolled in UK Biobank. Estimates were derived using data from the UK Biobank (app #76538).

## Data and computing code availability

Template code generated to run analyses outlined in this article are available on GitHub (https://github.com/gracemarionpower/SMM-MR/)

## Acknowledgements

We would like to thank the UK Biobank study and all participants who contributed to it, as well as the authors of all the GWAS who made their summary statistics available for the benefit of this work. This research has been conducted using the UK Biobank Resource under Application Number 76538. We would also like to thank Dr Joy Shi from the CAUSALab and Department of Epidemiology at Harvard T.H. Chan School of Public Health Cambridge, Massachusetts, United States for her insightful comments and intellectual input on this paper.

