## Supplementary Material for "A structural mean modelling Mendelian randomization approach to investigate the lifecourse effect of adiposity: applied and methodological considerations"

**Supplementary Table 1A. The full set of genetic variants strongly associated with childhood adiposity**
[Excel format]

**Supplementary Table 1B. The full set of genetic variants strongly associated with adulthood adiposity**

[Excel format]

**Supplementary Table 1C. The genetic variants strongly associated with childhood adiposity at genome wide significance and not adulthood adiposity (exclude adult SNPs at P ≤ 5×10^-8^)**

[Excel format]

**Supplementary Table 1D. The genetic variants strongly associated with childhood adiposity at genome wide significance and not adulthood adiposity (exclude adult SNPs at P ≤ 5×10^-8^) in females**

[Excel format]

**Supplementary Table 1E. The genetic variants strongly associated with adulthood adiposity at genome wide significance and not childhood adiposity (exclude child SNPs at P ≤ 5×10^-8^)**

[Excel format]

**Supplementary Table 1F. The genetic variants strongly associated with adulthood adiposity at genome wide significance and not childhood adiposity (exclude child SNPs at P ≤ 5×10^-8^) in females**

[Excel format]

**Supplementary Table 1G. The genetic variants strongly associated with childhood adiposity at genome wide significance and not adulthood adiposity (exclude adult SNPs at P ≤ 0.05 with Bonferroni correction*)**

[Excel format]

**Supplementary Table 1H. The genetic variants strongly associated with childhood adiposity at genome wide significance and not adulthood adiposity (exclude adult SNPs at P ≤ 0.05 with Bonferroni correction*) in females**

[Excel format]

**Supplementary Table 1I. The genetic variants strongly associated with adulthood adiposity at genome wide significance and not childhood adiposity (exclude child SNPs at P ≤ 0.05 with Bonferroni correction**)**

[Excel format]

**Supplementary Table 1J. The genetic variants strongly associated with adulthood adiposity at genome wide significance and not childhood adiposity (exclude child SNPs at P ≤ 0.05 with Bonferroni correction**) in females**

[Excel format]

**Supplementary Table 1K. The genetic variants strongly associated with childhood adiposity at genome wide significance and not adulthood adiposity (exclude adult SNPs at P ≤ 0.05)**

[Excel format]

**Supplementary Table 1L. The genetic variants strongly associated with childhood adiposity at genome wide significance and not adulthood adiposity (exclude adult SNPs at P ≤ 0.05) in females**

[Excel format]

**Supplementary Table 1M. The genetic variants strongly associated with adulthood adiposity at genome wide significance and not childhood adiposity (exclude child SNPs at P ≤ 0.05)**

[Excel format]

**Supplementary Table 1N. The genetic variants strongly associated with adulthood adiposity at genome wide significance and not childhood adiposity (exclude child SNPs at P ≤ 0.05) in females**
[Excel format]

* Genetic variants strongly associated with adulthood adiposity at genome wide significance and not childhood adiposity (exclude child SNPs at P ≤ 0.05) were counted. We then divided 0.05 by this number to generate the Bonferroni corrected P value.

** Genetic variants strongly associated with childhood adiposity at genome wide significance and not adulthood adiposity (exclude adult SNPs at P ≤ 0.05) were counted. We then divided 0.05 by this number to generate the Bonferroni corrected P value.

**Supplementary Table 2A. Univariable and multivariable Mendelian randomization (MR) analyses using structural mean modelling (SMM) for child and adult adiposity on cardiovascular disease (CVD) - using UKB outcome data**

| Exposure | Outcome | Risk difference | Standard error | P value | Sample size | MR |
| --- | --- | --- | --- | --- | --- | --- |
| Child associated SNPs* - regardless of adult SNP association | CVD | 0.021 | 0.004 | 1.75E-08 | 452960 | Univariable MR |
| Adult associated SNPs* - regardless of child SNP association | CVD | 0.062 | 0.003 | <2E-16 | 452960 | Univariable MR |
| Child associated SNPs* - regardless of adult SNP association, conditioning on adult associated SNPs | CVD | -0.024 | 0.005 | 3.92E-06 | 452960 | Multivariable MR |
| Adult associated SNPs* - regardless of child SNP association, conditioning on child associated SNPs | CVD | 0.074 | 0.004 | <2E-16 | 452960 | Multivariable MR |
| Child associated SNPs* - exclude adult associated SNPs at P ≤ 5*10^-8^ (genome-wide significance) | CVD | 0.004 | 0.006 | 0.419 | 452960 | Univariable MR |
| Adult associated SNPs* - exclude child associated SNPs at P ≤ 5*10^-8^ (genome-wide significance) | CVD | 0.067 | 0.004 | <2E-16 | 452960 | Univariable MR |
| Child associated SNPs* - exclude adult associated SNPs at P ≤ 0.05 with Bonferroni correction | CVD | 0.002 | 0.007 | 0.801 | 452960 | Univariable MR |
| Adult associated SNPs* - exclude child associated SNPs at P ≤ 0.05 with Bonferroni correction | CVD | 0.065 | 0.004 | <2E-16 | 452960 | Univariable MR |
| Child associated SNPs* - exclude adult associated SNPs at P ≤ 0.05 | CVD | -0.002 | 0.010 | 0.809 | 452960 | Univariable MR |
| Adult associated SNPs* - exclude child associated SNPs at P ≤ 0.05 | CVD | 0.070 | 0.005 | <2E-16 | 452960 | Univariable MR |

*Associated SNPs refers to strongly associated at genome wide significance (P≤ 5×10^-8^)

**Supplementary Table 2B. Univariable and multivariable Mendelian randomization analyses using structural mean models for child and adult adiposity on type 2 diabetes (T2D) - using UKB outcome data**

| Exposure | Outcome | Risk difference | Standard error | P value | Sample size | MR |
| --- | --- | --- | --- | --- | --- | --- |
| Child associated SNPs* - regardless of adult SNP association | T2D | 0.049 | 0.003 | <2E-16 | 452960 | Univariable MR |
| Adult associated SNPs* - regardless of child SNP association | T2D | 0.107 | 0.003 | <2E-16 | 452960 | Univariable MR |
| Child associated SNPs* - regardless of adult SNP association, conditioning on adult associated SNPs | T2D | -0.024 | 0.004 | 5.39E-08 | 452960 | Multivariable MR |
| Adult associated SNPs* - regardless of child SNP association, conditioning on child associated SNPs | T2D | 0.118 | 0.004 | <2E-16 | 452960 | Multivariable MR |
| Child associated SNPs* - exclude adult associated SNPs at P ≤ 5*10^-8^ (genome-wide significance) | T2D | 0.018 | 0.005 | 1.64E-04 | 452960 | Univariable MR |
| Adult associated SNPs* - exclude child associated SNPs at P ≤ 5*10^-8^ (genome-wide significance) | T2D | 0.109 | 0.003 | <2E-16 | 452960 | Univariable MR |
| Child associated SNPs* - exclude adult associated SNPs at P ≤ 0.05 with Bonferroni correction | T2D | -0.004 | 0.006 | 4.69E-01 | 452960 | Univariable MR |
| Adult associated SNPs* - exclude child associated SNPs at P ≤ 0.05 with Bonferroni correction | T2D | 0.108 | 0.003 | <2E-16 | 452960 | Univariable MR |
| Child associated SNPs* - exclude adult associated SNPs at P ≤ 0.05 | T2D | -0.008 | 0.008 | 0.364 | 452960 | Univariable MR |
| Adult associated SNPs* - exclude child associated SNPs at P ≤ 0.05 | T2D | 0.115 | 0.004 | <2E-16 | 452960 | Univariable MR |

*Associated SNPs refers to strongly associated at genome wide significance (P≤ 5×10^-8^)

**Supplementary Table 2C. Univariable and multivariable Mendelian randomization analyses using structural mean models for child and adult adiposity on breast cancer - using UKB outcome data**

| Exposure | Outcome | Risk difference | Standard error | P value | Sample size | MR | Population |
| --- | --- | --- | --- | --- | --- | --- | --- |
| Child associated SNPs* - regardless of adult SNP association | Breast cancer | -0.032 | 0.005 | 6.72E-12 | 246404 | Univariable MR | Female |
| Adult associated SNPs* - regardless of child SNP association | Breast cancer | -0.008 | 0.004 | 0.053 | 246404 | Univariable MR | Female |
| Child associated SNPs* - regardless of adult SNP association, conditioning on adult associated SNPs | Breast cancer | -0.040 | 0.007 | 8.27E-10 | 246404 | Multivariable MR | Female |
| Adult associated SNPs* - regardless of child SNP association, conditioning on child associated SNPs | Breast cancer | 0.013 | 0.006 | 0.03 | 246404 | Multivariable MR | Female |
| Child associated SNPs* - exclude adult associated SNPs at P ≤ 5*10^-8^ (genome-wide significance) | Breast cancer | -0.029 | 0.007 | 3.42E-05 | 246404 | Univariable MR | Female |
| Adult associated SNPs* - exclude child associated SNPs at P ≤ 5*10^-8^ (genome-wide significance) | Breast cancer | 0.006 | 0.005 | 1.83E-01 | 246404 | Univariable MR | Female |
| Child associated SNPs* - exclude adult associated SNPs at P ≤ 0.05 with Bonferroni correction | Breast cancer | -0.029 | 0.009 | 9.22E-04 | 246407 | Univariable MR | Female |
| Adult associated SNPs* - exclude child associated SNPs at P ≤ 0.05 with Bonferroni correction | Breast cancer | 0.014 | 0.005 | 0.0106 | 246408 | Univariable MR | Female |
| Child associated SNPs* - exclude adult associated SNPs at P ≤ 0.05 | Breast cancer | -0.035 | 0.012 | 2.93E-03 | 246405 | Univariable MR | Female |
| Adult associated SNPs* - exclude child associated SNPs at P ≤ 0.05 | Breast cancer | 0.017 | 0.006 | 7.15E-03 | 246406 | Univariable MR | Female |

*Associated SNPs refers to strongly associated at genome wide significance (P≤ 5×10^-8^)

**Supplementary Table 3A. Univariable and multivariable two-sample Mendelian randomization (MR) analyses using inverse probability weighting for child and adult adiposity on cardiovascular disease (CVD) - using UKB outcome data**

| Exposure | Outcome | nSNP | Risk difference | Standard error | P value | Sample size | MR |
| --- | --- | --- | --- | --- | --- | --- | --- |
| Child associated SNPs* - regardless of adult SNP association | CVD | 313 | 0.022 | 0.005 | 1.28E-06 | 452960 | Univariable MR |
| Adult associated SNPs* - regardless of child SNP association | CVD | 580 | 0.062 | 0.004 | 1.24E-52 | 452960 | Univariable MR |
| Child associated SNPs* - regardless of adult SNP association, conditioning on adult associated SNPs | CVD | 267 | -0.021 | 0.006 | 1.18E-03 | 452960 | Multivariable MR |
| Adult associated SNPs* - regardless of child SNP association, conditioning on child associated SNPs | CVD | 535 | 0.070 | 0.006 | 9.59E-37 | 452960 | Multivariable MR |
| Child associated SNPs* - exclude adult associated SNPs at P ≤ 5*10^-8^ (genome-wide significance) | CVD | 180 | 0.007 | 0.006 | 2.93E-01 | 452960 | Univariable MR |
| Adult associated SNPs* - exclude child associated SNPs at P ≤ 5*10^-8^ (genome-wide significance) | CVD | 439 | 0.069 | 0.005 | 5.35E-43 | 452960 | Univariable MR |
| Child associated SNPs* - exclude adult associated SNPs at P ≤ 0.05 with Bonferroni correction | CVD | 106 | 0.006 | 0.010 | 5.43E-01 | 452960 | Univariable MR |
| Adult associated SNPs* - exclude child associated SNPs at P ≤ 0.05 with Bonferroni correction | CVD | 365 | 0.067 | 0.006 | 8.80E-32 | 452960 | Univariable MR |
| Child associated SNPs* - exclude adult associated SNPs at P ≤ 0.05 | CVD | 55 | -0.005 | 0.014 | 7.51E-01 | 452960 | Univariable MR |
| Adult associated SNPs* - exclude child associated SNPs at P ≤ 0.05 | CVD | 218 | 0.071 | 0.008 | 2.88E-21 | 452960 | Univariable MR |

*Associated SNPs refers to strongly associated at genome wide significance (P≤ 5×10^-8^)

**Supplementary Table 3B. Univariable and multivariable two-sample Mendelian randomization (MR) analyses using inverse probability weighting for child and adult adiposity on type 2 diabetes (T2D) - using UKB outcome data**

| Exposure | Outcome | nSNP | Risk difference | Standard error | P value | Sample size | MR |
| --- | --- | --- | --- | --- | --- | --- | --- |
| Child associated SNPs* - regardless of adult SNP association | T2D | 313 | 0.055 | 0.005 | 2.09E-29 | 452960 | Univariable MR |
| Adult associated SNPs* - regardless of child SNP association | T2D | 580 | 0.113 | 0.005 | 1.39E-134 | 452960 | Univariable MR |
| Child associated SNPs* - regardless of adult SNP association, conditioning on adult associated SNPs | T2D | 267 | -0.022 | 0.007 | 1.87E-03 | 452960 | Multivariable MR |
| Adult associated SNPs* - regardless of child SNP association, conditioning on child associated SNPs | T2D | 535 | 0.123 | 0.006 | 4.20E-91 | 452960 | Multivariable MR |
| Child associated SNPs* - exclude adult associated SNPs at P ≤ 5*10^-8^ (genome-wide significance) | T2D | 180 | 0.021 | 0.007 | 2.60E-03 | 452960 | Univariable MR |
| Adult associated SNPs* - exclude child associated SNPs at P ≤ 5*10^-8^ (genome-wide significance) | T2D | 439 | 0.112 | 0.006 | 1.64E-77 | 452960 | Univariable MR |
| Child associated SNPs* - exclude adult associated SNPs at P ≤ 0.05 with Bonferroni correction | T2D | 106 | -0.005 | 0.010 | 6.33E-01 | 452960 | Univariable MR |
| Adult associated SNPs* - exclude child associated SNPs at P ≤ 0.05 with Bonferroni correction | T2D | 365 | 0.111 | 0.007 | 6.61E-56 | 452960 | Univariable MR |
| Child associated SNPs* - exclude adult associated SNPs at P ≤ 0.05 | T2D | 55 | -0.011 | 0.016 | 4.72E-01 | 452960 | Univariable MR |
| Adult associated SNPs* - exclude child associated SNPs at P ≤ 0.05 | T2D | 218 | 0.117 | 0.006 | 5.58E-80 | 452960 | Univariable MR |

*Associated SNPs refers to strongly associated at genome wide significance (P≤ 5×10^-8^)

**Supplementary Table 3C. Univariable and multivariable two-sample Mendelian randomization (MR) analyses using inverse probability weighting for child and adult adiposity on breast cancer - using UKB outcome data**

| Exposure | Outcome | nSNP | Risk difference | Standard error | P value | Sample size | MR | Population |
| --- | --- | --- | --- | --- | --- | --- | --- | --- |
| Child associated SNPs* - regardless of adult SNP association | Breast cancer | 142 | -0.018 | 0.003 | 1.34E-09 | 246404 | Univariable MR | Female |
| Adult associated SNPs* - regardless of child SNP association | Breast cancer | 221 | -0.004 | 0.003 | 2.06E-01 | 246404 | Univariable MR | Female |
| Child associated SNPs* - regardless of adult SNP association, conditioning on adult associated SNPs | Breast cancer | 132 | -0.023 | 0.004 | 1.08E-09 | 246404 | Multivariable MR | Female |
| Adult associated SNPs* - regardless of child SNP association, conditioning on child associated SNPs | Breast cancer | 202 | 0.009 | 0.004 | 1.58E-02 | 246404 | Multivariable MR | Female |
| Child associated SNPs* - exclude adult associated SNPs at P ≤ 5*10^-8^ (genome-wide significance) | Breast cancer | 95 | -0.015 | 0.004 | 4.49E-05 | 246404 | Univariable MR | Female |
| Adult associated SNPs* - exclude child associated SNPs at P ≤ 5*10^-8^ (genome-wide significance) | Breast cancer | 172 | 0.004 | 0.003 | 2.27E-01 | 246406 | Univariable MR | Female |
| Child associated SNPs* - exclude adult associated SNPs at P ≤ 0.05 with Bonferroni correction | Breast cancer | 50 | -0.015 | 0.006 | 5.23E-03 | 246405 | Univariable MR | Female |
| Adult associated SNPs* - exclude child associated SNPs at P ≤ 0.05 with Bonferroni correction | Breast cancer | 147 | 0.008 | 0.003 | 2.04E-02 | 246407 | Univariable MR | Female |
| Child associated SNPs* - exclude adult associated SNPs at P ≤ 0.05 | Breast cancer | 28 | -0.018 | 0.006 | 4.98E-03 | 246405 | Univariable MR | Female |
| Adult associated SNPs* - exclude child associated SNPs at P ≤ 0.05 | Breast cancer | 102 | 0.010 | 0.004 | 2.41E-02 | 246408 | Univariable MR | Female |

*Associated SNPs refers to strongly associated at genome wide significance (P≤ 5×10^-8^)

**Supplementary Table 4A. Univariable and multivariable two-sample Mendelian randomization (MR) analyses using inverse probability weighting for child and adult adiposity on cardiovascular (CVD) – using large scale consortium data**

| Exposure | Outcome | nSNP | Risk difference | Standard error | P value | Sample size | MR |
| --- | --- | --- | --- | --- | --- | --- | --- |
| Child associated SNPs* - regardless of adult SNP association | CVD | 245 | 0.418 | 0.065 | 9.57E-11 | 452960 | Univariable MR |
| Adult associated SNPs* - regardless of child SNP association | CVD | 463 | 0.554 | 0.056 | 6.71E-23 | 452960 | Univariable MR |
| Child associated SNPs* - regardless of adult SNP association, conditioning on adult associated SNPs | CVD | 169 | -0.154 | 0.226 | 4.96E-01 | 452960 | Multivariable MR |
| Adult associated SNPs* - regardless of child SNP association, conditioning on child associated SNPs | CVD | 318 | 1.126 | 0.195 | 7.65E-09 | 452960 | Multivariable MR |
| Child associated SNPs* - exclude adult associated SNPs at P ≤ 5*10^-8^ (genome-wide significance) | CVD | 148 | 0.276 | 0.095 | 3.82E-03 | 452960 | Univariable MR |
| Adult associated SNPs* - exclude child associated SNPs at P ≤ 5*10^-8^ (genome-wide significance) | CVD | 362 | 0.556 | 0.074 | 6.36E-14 | 452960 | Univariable MR |
| Child associated SNPs* - exclude adult associated SNPs at P ≤ 0.05 with Bonferroni correction | CVD | 85 | 0.243 | 0.135 | 7.21E-02 | 452960 | Univariable MR |
| Adult associated SNPs* - exclude child associated SNPs at P ≤ 0.05 with Bonferroni correction | CVD | 297 | 0.502 | 0.087 | 6.59E-09 | 452960 | Univariable MR |
| Child associated SNPs* - exclude adult associated SNPs at P ≤ 0.05 | CVD | 47 | -0.027 | 0.181 | 8.83E-01 | 452960 | Univariable MR |
| Adult associated SNPs* - exclude child associated SNPs at P ≤ 0.05 | CVD | 172 | 0.548 | 0.116 | 2.40E-06 | 452960 | Univariable MR |

*Associated SNPs refers to strongly associated at genome wide significance (P≤ 5×10^-8^)

**Supplementary Table 4B. Univariable and multivariable two-sample Mendelian randomization (MR) analyses using inverse probability weighting for child and adult adiposity on type 2 diabetes (T2D) – using large scale consortium data**

| Exposure | Outcome | nSNP | Risk difference | Standard error | P value | Sample size | MR |
| --- | --- | --- | --- | --- | --- | --- | --- |
| Child associated SNPs* - regardless of adult SNP association | T2D | 163 | 0.738 | 0.157 | 2.55E-06 | 452960 | Univariable MR |
| Adult associated SNPs* - regardless of child SNP association | T2D | 286 | 1.158 | 0.163 | 1.05E-12 | 452960 | Univariable MR |
| Child associated SNPs* - regardless of adult SNP association, conditioning on adult associated SNPs | T2D | 169 | -0.154 | 0.226 | 4.96E-01 | 452960 | Multivariable MR |
| Adult associated SNPs* - regardless of child SNP association, conditioning on child associated SNPs | T2D | 318 | 1.126 | 0.195 | 7.65E-09 | 452960 | Multivariable MR |
| Child associated SNPs* - exclude adult associated SNPs at P ≤ 5*10^-8^ (genome-wide significance) | T2D | 114 | 0.460 | 0.224 | 3.97E-02 | 452960 | Univariable MR |
| Adult associated SNPs* - exclude child associated SNPs at P ≤ 5*10^-8^ (genome-wide significance) | T2D | 254 | 0.839 | 0.194 | 1.49E-05 | 452960 | Univariable MR |
| Child associated SNPs* - exclude adult associated SNPs at P ≤ 0.05 with Bonferroni correction | T2D | 67 | 0.302 | 0.346 | 3.82E-01 | 452960 | Univariable MR |
| Adult associated SNPs* - exclude child associated SNPs at P ≤ 0.05 with Bonferroni correction | T2D | 208 | 0.799 | 0.231 | 5.51E-04 | 452960 | Univariable MR |
| Child associated SNPs* - exclude adult associated SNPs at P ≤ 0.05 | T2D | 35 | 0.115 | 0.465 | 8.05E-01 | 452960 | Univariable MR |
| Adult associated SNPs* - exclude child associated SNPs at P ≤ 0.05 | T2D | 120 | 1.071 | 0.222 | 1.35E-06 | 452960 | Univariable MR |

*Associated SNPs refers to strongly associated at genome wide significance (P≤ 5×10^-8^)

**Supplementary Table 4C. Univariable and multivariable two-sample Mendelian randomization (MR) analyses using inverse probability weighting for child and adult adiposity on breast cancer – using large scale consortium data**

| Exposure | Outcome | nSNP | Risk difference | Standard error | P value | Sample size | MR | Population |
| --- | --- | --- | --- | --- | --- | --- | --- | --- |
| Child associated SNPs* - regardless of adult SNP association | Breast cancer | 114 | -0.460 | 0.072 | 1.63E-10 | 246404 | Univariable MR | Female |
| Adult associated SNPs* - regardless of child SNP association | Breast cancer | 167 | -0.245 | 0.065 | 1.80E-04 | 246404 | Univariable MR | Female |
| Child associated SNPs* - regardless of adult SNP association, conditioning on adult associated SNPs | Breast cancer | 12 | -0.631 | 0.187 | 7.29E-04 | 246404 | Multivariable MR | Female |
| Adult associated SNPs* - regardless of child SNP association, conditioning on child associated SNPs | Breast cancer | 143 | 0.131 | 0.094 | 1.66E-01 | 246404 | Multivariable MR | Female |
| Child associated SNPs* - exclude adult associated SNPs at P ≤ 5*10^-8^ (genome-wide significance) | Breast cancer | 67 | -0.328 | 0.092 | 3.75E-04 | 246404 | Univariable MR | Female |
| Adult associated SNPs* - exclude child associated SNPs at P ≤ 5*10^-8^ (genome-wide significance) | Breast cancer | 110 | 0.039 | 0.074 | 6.03E-01 | 246404 | Univariable MR | Female |
| Child associated SNPs* - exclude adult associated SNPs at P ≤ 0.05 with Bonferroni correction | Breast cancer | 34 | -0.237 | 0.103 | 2.13E-02 | 246404 | Univariable MR | Female |
| Adult associated SNPs* - exclude child associated SNPs at P ≤ 0.05 with Bonferroni correction | Breast cancer | 97 | 0.075 | 0.080 | 3.51E-01 | 246406 | Univariable MR | Female |
| Child associated SNPs* - exclude adult associated SNPs at P ≤ 0.05 | Breast cancer | 18 | -0.297 | 0.158 | 6.03E-02 | 246407 | Univariable MR | Female |
| Adult associated SNPs* - exclude child associated SNPs at P ≤ 0.05 | Breast cancer | 68 | 0.089 | 0.094 | 3.44E-01 | 246408 | Univariable MR | Female |

*Associated SNPs refers to strongly associated at genome wide significance (P≤ 5×10^-8^)

**Supplementary Figure 5. Causal risk difference estimates from univariable and multivariable MR using inverse variance weighted models (IVW) for childhood and adult adiposity period effects on the outcome measures listed using large scale consortia data referenced in manuscript.** (A) Childhood and adult adiposity period and lifetime effects on cardiovascular disease (CVD). (B) Childhood and adult adiposity period and lifetime effects on Type 2 diabetes (T2D). (C) Childhood and adult adiposity period and lifetime effects on breast cancer. Child/adult only/only SNPs indicates genetic variants strongly associated with childhood and adult adiposity (using P < 5x10^-8^).

A

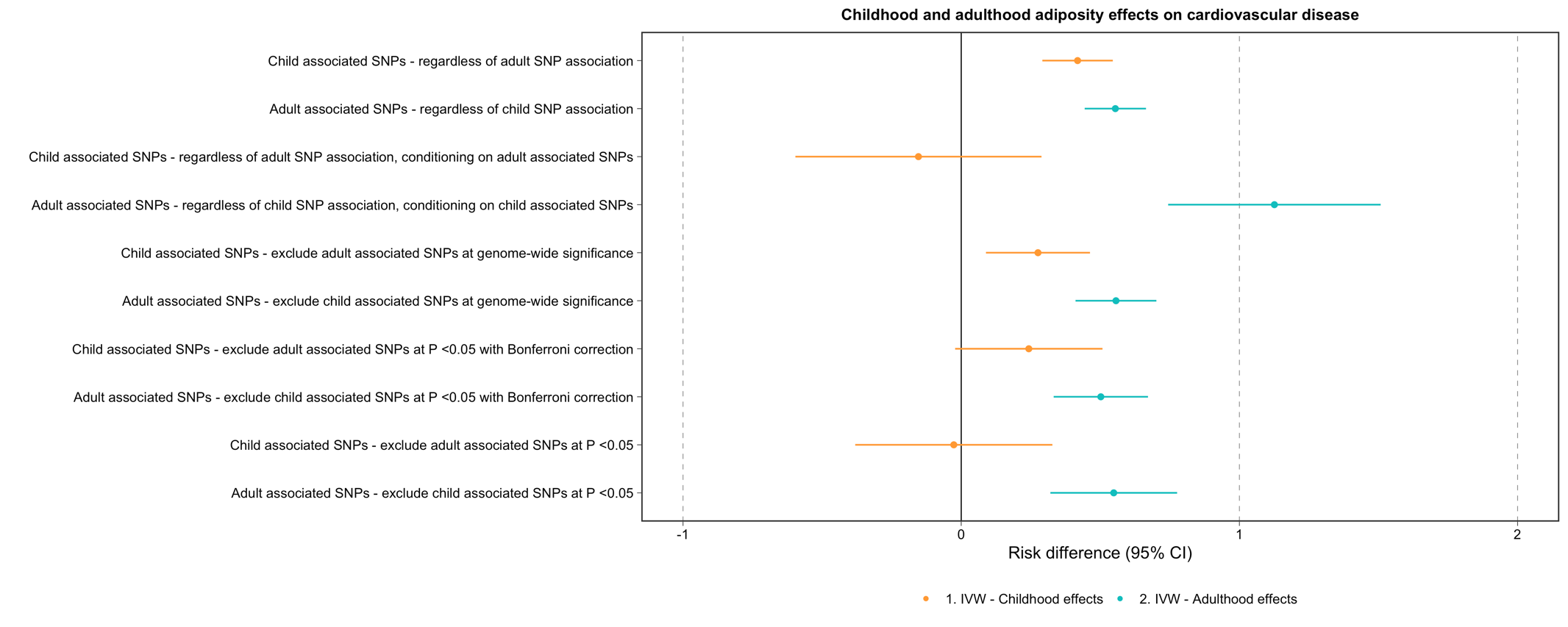

B

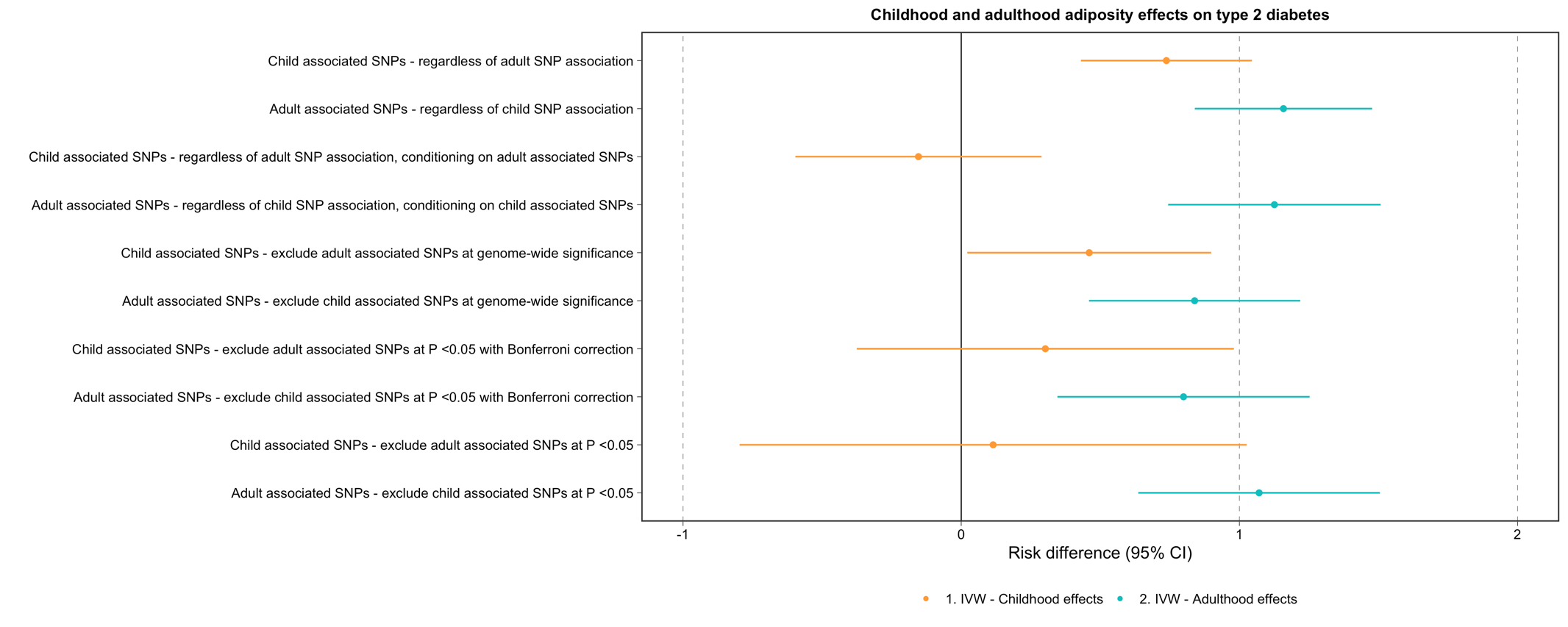

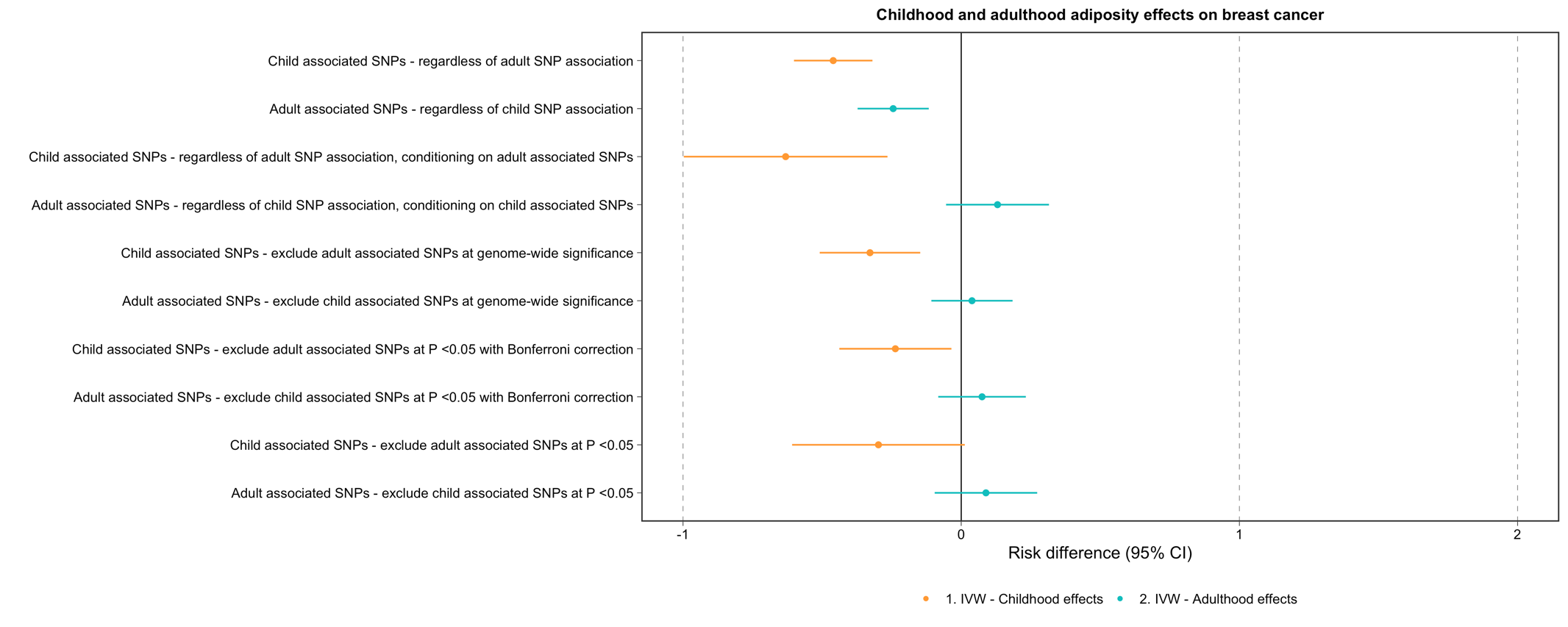

C
